# Identification and characterization of human GDF15 knockouts

**DOI:** 10.1101/2024.03.14.24303793

**Authors:** Allan M. Gurtan, Shareef Khalid, Christopher Koch, Maleeha Zaman Khan, Lindsey B. Lamarche, Elizabeth Dolan, Ana M. Carrion, Richard Zessis, Matthew E. Clement, Zhiping Chen, Loren D. Lindsley, Yu-Hsin Chiu, Ryan S. Streeper, Daniel P. Denning, Allison B. Goldfine, Brian Doyon, Ali Abbasi, Jennifer L. Harrow, Kazuhisa Tsunoyama, Makoto Asaumi, Ikuyo Kou, Alan R. Shuldiner, Juan L. Rodriguez-Flores, Asif Rasheed, Muhammad Jahanzaib, Muhammad Rehan Mian, Muhammad Bilal Liaqat, Syed Shahzaib Raza, Riffat Sultana, Anjum Jalal, Muhammad Hamid Saeed, Shahid Abbas, Fazal Rehman Memon, Muhammad Ishaq, John E. Dominy, Danish Saleheen

## Abstract

Growth differentiation factor 15 (GDF15) is a secreted protein that regulates food intake, body weight, and stress responses in pre-clinical models^1^. The physiological function of GDF15 in humans remains unclear. Pharmacologically, GDF15 agonism in humans caused nausea without accompanying weight loss^2^, and the effect of GDF15 antagonism is being tested in clinical trials to treat cachexia and anorexia. Human genetics point to a role for GDF15 in hyperemesis gravidarum, but the safety or impact of complete GDF15 loss, particularly during pregnancy, is unknown^3–7^. Here, we characterize GDF15 loss-of-function carriers (LOF), including 5 homozygous null carriers (“knockouts”) from 75,018 participants enrolled in the Pakistan Genomic Resource (PGR)^8,9^. We tested for the association of GDF15 LOF with 97 quantitative traits and binary outcomes. Further, 3 additional knockouts and 59 heterozygous carriers were identified in recall-by-genotype (RBG) studies accompanied by recruitment of family members. GDF15 knockouts ranged in age from 31 to 75 years, were fertile, and showed no consistent overt metabolic dysfunction. Collectively, our data indicate that (i) complete GDF15 loss is compatible with life and fertility, (ii) chronic therapeutic inhibition of GDF15 may be tolerated, and (iii) GDF15 modulation may not significantly impact body weight or metabolic syndrome.

## Main

Selective and safe control of the interconnected pathways of appetite, satiety, nausea, and emesis has become a therapeutic objective. Hundreds of millions of patients worldwide suffer from a range of disorders that could be treated with novel medicines that impact these processes^10,11^. Therapies that suppress appetite or stimulate satiety may help treat obesity and type 2 diabetes by curbing food intake and reducing weight. GLP1 receptor agonists, for example, reduce body weight in humans through appetite suppression and improved metabolic control^10^. Conversely, therapeutics that increase appetite or suppress nausea and emesis may treat disorders such as anorexia, cachexia, hyperemesis gravidarum, and associated sequelae^11^.

The growth differentiation factor 15 (GDF15) protein pathway was identified in pre-clinical models as a potential therapeutic target for appetite regulation^1^. This secreted protein is expressed in multiple tissues and is inducible under stress conditions and pregnancy. Its receptor, GFRAL, is expressed in the area postrema, an area of the brain containing circuitry governing hunger and emesis responses^12–14^. Pharmacological administration of GDF15 causes weight loss in wild-type mice through reduction of food intake^15–17^. This effect is lost in mice that are GFRAL knockout (KO) or have been treated with a GDF15 antagonist antibody^12–14^. Body weight is lower in GDF15-expressing transgenic mice^15,16^ and, conversely, higher in GDF15 KO^18^ and GFRAL KO^12–14^ mice. In mice treated with prostate cancer xenografts, elevated GDF15 concentrations are associated with anorexia-cachexia, and this effect is reversed by a GDF15-blocking antibody^15^. In spontaneously obese cynomolgus macaques, exogenous administration of GDF15 results in concomitant reduction of food intake and body weight^12^. Thus, in multiple pre-clinical models, modulation of GDF15 impacts body weight: activation or inhibition of the GDF15-GFRAL pathway respectively reduces or increases food intake and body weight^1,12–17^.

In humans, GDF15 is associated with health conditions that impact food intake, appetite, and body weight. During the first two trimesters of human pregnancy, a period where many women report higher incidence of nausea and emesis, *GDF15* is highly expressed by trophoblast cells of the placenta^7^. Genome-wide association studies (GWAS) have linked common genetic polymorphisms at the *GDF15* and *GFRAL* loci with hyperemesis gravidarum (HG)^4–7^, a condition that leads to severe nausea and emesis during pregnancy and can require hospitalization. Notably, a recent study suggests that mothers with germline *GDF15* variants causing partial loss of GDF15 expression are at an elevated risk for hyperemesis gravidarum if their fetus has a genotype associated with higher GDF15 secretion^3^. Beyond pregnancy, elevated plasma concentrations of GDF15 correlate with cachexia and are predictive of mortality across numerous diseases, including cancer, chronic renal disease, and cardiovascular disease^1,19^. Consistent with observations in preclinical models, human biomarker and genetic data point to a role for GDF15 signaling in food intake, appetite, and nausea/emesis. On the basis of these data, numerous efforts have been initiated to activate or inhibit the GDF15 pathway for treatment of metabolic conditions^20^.

An understanding of the phenotypic impact of complete loss of GDF15, from gestation into adulthood and the natural reproductive cycle, would inform GDF15 therapeutic programs. Through whole exome sequencing and recall-by-genotype studies, we identified and characterized the largest known cohort to date of heterozygous GDF15 LOF carriers. Further, we report the discovery and characterization of naturally occurring homozygous GDF15 LOF (GDF15 “knockout” [KO]) humans.

### Functional validation of naturally occurring human GDF15 loss-of-function (LOF) variants

The Pakistan Genomic Resource (PGR) at the Center for Non-Communicable Diseases (CNCD) in Pakistan is one of the world’s largest exome biobanks of human homozygous LOF carriers (human “knockouts”)^8^. In the PGR, we identified a total of 13 LOF variants comprising protein truncating nonsense and frameshift substitutions in *GDF15*, with a cumulative allele frequency of 0.027% (Extended Data Table 1) that includes one predicted KO. Additionally, a review of missense variants that may impact function revealed a variant, Cys211Gly (C211G), in a position that is highly conserved across species and across GDNF family members (Figure 1A) and has been recently suggested to be LOF^3^. In PGR, we identified 123 heterozygous carriers and 4 homozygous carriers of C211G (Extended Data Table 1), with an allele frequency of 0.086%. Baseline characteristics such as anthropometric traits, biochemical tests and medical history for LOF and C211G carriers are shown in Extended Data Table 2.

**Figure 1.**
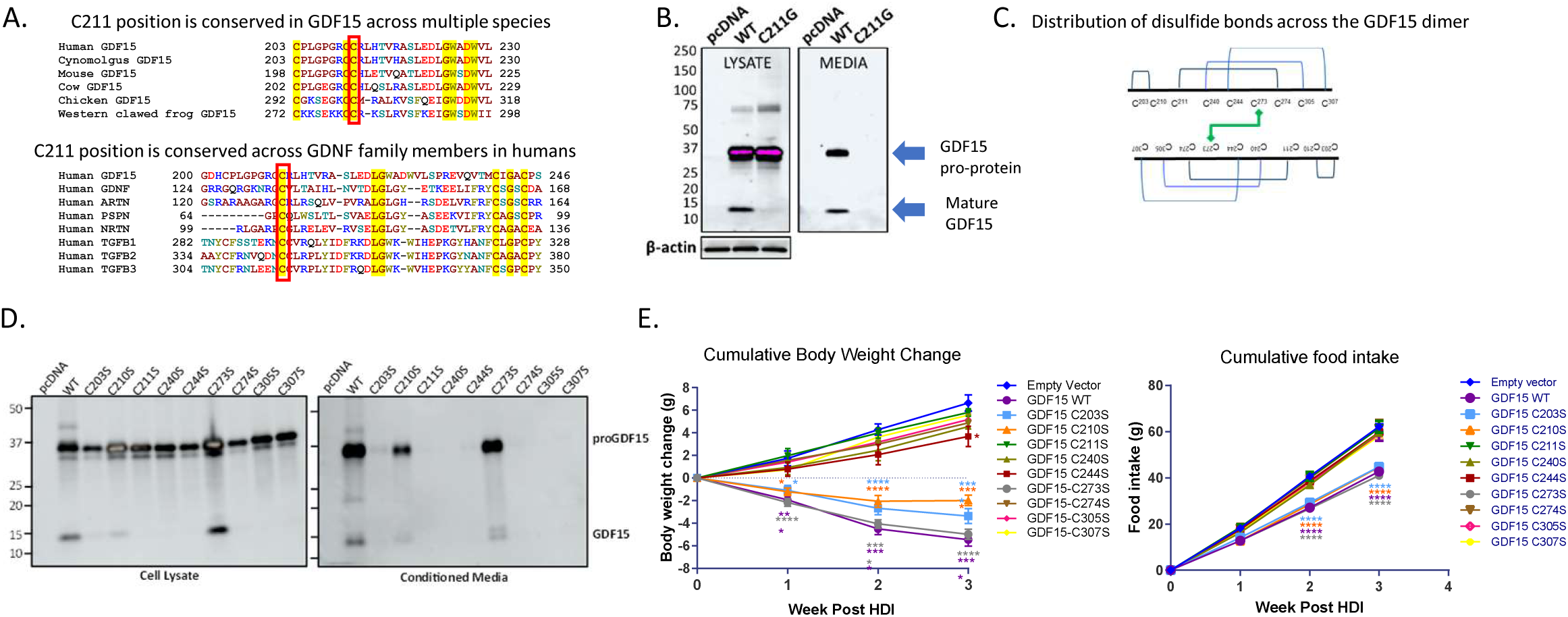
Cys211 is required for GDF15 secretion and activity. (A) Amino acid sequence alignment of GDF15 proteins across multiple vertebrates (top panel) and across paralogs in humans (bottom panel) shows strict conservation of C211 (boxed in red). (B) Western blot of human GDF15 variant C211G in transiently transfected HEK293 cells. A C-terminal APP tag was used for detection by western blot. (C) A schematic representation of the intra– and inter-molecular disulfide bonds present in the mature GDF15 dimer. (D) Identification by western blot of cysteine residues critical for GDF15 expression and processing. (E) Identification of cysteine residues critical for GDF15 activity *in vivo*, determined by measurement of body weight change (left panel) and cumulative food intake (right panel) in mice 14 days after hydrodynamic tail vein injection with the indicated GDF15 expression constructs. Mice were on a high fat diet for at least 8 weeks to induce obesity. N = 5-6 mice per treatment group. Statistical analyses conducted by two-way ANOVA with multiple comparison testing of each treatment vs the empty vector control. *p<0.05, ***P<0.001, ****P<0.0001 vs empty vector group.

**Table 1.** Distribution of baseline characteristics of recall-by-genotype participants per genotype (both C211G and LOF variant carriers and non-carriers).

|  |  | HomRR | HetRA | HomAA |  |  |
| --- | --- | --- | --- | --- | --- | --- |
|  | Count | median (IQR)<br>or % | median (IQR) or<br>% | median (IQR) or<br>% | P-val<br>(additive) | P-val<br>(recessive) |
| **Quantitative traits** |  |  |  |  |  |  |
| Age | 120:67:5 | 38 (28,50) | 37 (27,51.5) | 38 (32,64) |  |  |
| Gender (Female) | 78:33:1 | 65% | 49.3% | 20% |  |  |
| GDF15 plasma<br>concentration (ng/ml) | 62:41:4 | 623 (357,893) | 272 (140,662) | 8.6 (5.3,21.6) | 1.4e-7 | 7.4e-6 |
| BMI (kg/m<sup>2</sup>) | 111:61:5 | 25.2 (20.8,29.0) | 25.2 (21.6,28.8) | 29.0 (26.1,29.8) | 0.03 | 0.11 |
| Height (cm) | 118:67:5 | 156 (153,163) | 158 (152.5,167) | 162 (160,171) | 0.95 | 0.76 |
| Weight (kg) | 112:61:5 | 63.2 (51.0,<br>72.0) | 66.3 (56.0, 73.8) | 76.4 (76.0, 80.0) | 0.03 | 0.10 |
| WHR | 117:67:5 | 0.90 (0.86,0.93) | 0.94 (0.89,0.97) | 0.95 (0.92,0.95) | 0.06 | 0.91 |
| Waist (cm) | 117:67:5 | 87 (76,103) | 92 (83,105) | 96 (96,96) | 8.6e-3 | 0.21 |
| Hip (cm) | 117:67:5 | 97 (89,107) | 99 (91,112) | 104 (101,110) | 0.01 | 0.07 |
| BP systolic (mmHg) | 117:66:5 | 120 (113,131) | 120 (112,134) | 126 (125,130) | 0.86 | 0.74 |
| BP diastolic (mmHg) | 117:66:5 | 80 (75,89) | 80 (70,85) | 84 (80,85) | 0.14 | 0.78 |
| Heart rate (bpm) | 99:52:3 | 83 (76, 96) | 83 (74, 93) | 90 (83, 107) | 0.54 | 0.18 |
| Fat mass (kg) | 83:44:2 | 16 (9, 27) | 20 (12,29) | 23 (19, 27) | 0.03 | 0.97 |
| Fat free mass (kg) | 83:44:2 | 42 (36,46) | 44 (39,53) | 49 (48,51) | 0.40 | 0.82 |
| Cholesterol (mg/dl) | 120:67:5 | 155.5<br>(131.8,180) | 168.5 (141.9,198.9) | 184.1<br>(158.3,205.1) | 0.04 | 0.62 |
| Triglycerides (mg/dl) | 120:67:5 | 124.4<br>(89.2,168.3) | 143.5 (107.9,195.8) | 504<br>(225.3,652.3) | 0.12 | 0.05 |
| HDL-C (mg/dl) | 120:67:5 | 36 (31,42) | 36 (31,43.5) | 29 (22,32) | 0.82 | 0.03 |
| LDL-C (mg/dl) | 120:67:5 | 87 (71,108) | 97 (77,125) | 75 (46,99) | 0.40 | 0.04 |
| Creatinine (mg/dl) | 120:67:5 | 0.61 (0.55,0.86) | 0.75 (0.60,0.94) | 0.9 (0.57,0.9) | 0.09 | 0.18 |
| eGFR (mL/min/1.73m<sup>2</sup>) | 120:67:5 | 107 (85,139) | 104 (83,128) | 98 (82,123) | 0.16 | 0.17 |
| HbA1c (%) | 110:53:5 | 5.4 (5.1,5.7) | 5.5 (5.2,5.9) | 5.5 (5.4,6.3) | 0.46 | 0.78 |
| Glucose (mg/dl) | 120:67:5 | 83 (77,93) | 84 (73,106) | 81 (59,87) | 0.79 | 0.94 |
| AST (U/L) | 120:67:5 | 22 (18,26) | 23 (18,30) | 29 (26,29) | 0.18 | 0.30 |
| ALT (U/L) | 120:67:5 | 20 (13,26) | 19 (14,26) | 23 (22,30) | 0.21 | 0.34 |
| ALP (U/L) | 87:57:4 | 73 (60,103) | 92 (77,118) | 97 (92,103) | 0.20 | 0.94 |
| White Blood Cells (10<sup>9</sup> / L) | 89:45:2 | 6.5 (4.2,8.3) | 6.3 (3.0,8.1) | 6.2 (5.3,7.0) | 0.11 | 0.43 |
| Red Blood Cells (10<sup>12</sup> / L) | 89:45:2 | 4.7 (4.1,5.1) | 4.9 (4.4,5.4) | 5.1 (5.0,5.1) | 1.00 | 0.77 |
| Hemoglobin (g/dl) | 89:45:2 | 13.1 (11.8,14.3) | 13.2 (12.4,14.6) | 14.4 (13.7,15.0) | 0.93 | 0.34 |
| Hematocrit (%) | 89:45:2 | 39.4 (33.5,44.1) | 42.2 (38.4,46.1) | 47.1 (46.3,47.8) | 0.72 | 0.34 |
| MCV (fl) | 89:45:2 | 84 (77,89) | 86 (82,90) | 93 (92,9) | 0.76 | 0.23 |
| MCH (pg) | 89:45:2 | 28 (26,30) | 27 (26,29) | 28 (27,29) | 0.73 | 0.52 |
| MCHC (g/dl) | 89:45:2 | 31.5 (30.2,35.2) | 31.3 (29.7,32.3) | 30.45 (29.6,31.3) | 0.91 | 0.98 |
| Platelets (10<sup>9</sup> / L) | 89:45:2 | 330 (243,460) | 287 (188,360) | 240 (232,249) | 0.30 | 0.88 |
| Neutrophils (%) | 89:45:2 | 40.2 (25.9,50.1) | 36.6 (21.4,55.0) | 37.6(29.8,45.4) | 0.45 | 0.84 |
| Lymphocytes (%) | 89:45:2 | 41.8 (33.9,50.2) | 43.2 (33.8,51.1) | 44.7 (39.7,49.6) | 0.83 | 0.78 |
| Monocytes (%) | 89:45:2 | 8.2 (6.3,11) | 9.5 (7.5,12.4) | 10.2 (10,10.4) | 0.39 | 0.83 |
| Eosinophils (%) | 89:45:2 | 4.1 (2.3,6) | 3.5 (2.7,6) | 3.7 (2.7,4.7) | 0.12 | 0.43 |
| Basophils (%) | 89:45:2 | 2.2 (0.8,9) | 2.5 (0.7,6.5) | 3.9 (2.2,5.6) | 0.06 | 0.33 |
| Urea (mg/dl) | 87:57:4 | 23 (19.5,28) | 24 (18,31) | 29.5 (26.3,32.3) | 0.15 | 0.28 |
| BIT (mg/dl) | 87:57:4 | 0.26 (0.14,0.37) | 0.24 (0.15,0.34) | 0.26 (0.22,0.29) | 0.18 | 0.85 |
| Ferritin (ng/ml) | 87:57:4 | 57.5<br>(22.1,101.4) | 63.5 (22.8,129.1) | 59.3 (53.3,97.4) | 0.82 | 0.93 |
| Calcium (mg/dl) | 87:57:4 | 9.6 (8.8,10.6) | 9.3 (8.9,10.2) | 10.7 (9.7,11.7) | 0.93 | 0.30 |
| Sodium (mmol/L) | 87:57:4 | 145 (140,152) | 144 (140,151) | 154 (150,163) | 0.86 | 0.22 |
| Potassium (mmol/L) | 87:57:4 | 4.6 (4.2,5.2) | 4.7 (4.3,5.3) | 5.6 (4.9,6.5) | 0.29 | 0.05 |
| Urinary microalbumin (mg/dl) | 107:65:5 | 0.56 (0.15,1.93) | 0.48 (0.19,1.62) | 0.85 (0.42,1.52) | 0.23 | 0.69 |
| Leptin plasma concentration (ng/ml) | 59:40:4 | 5.2 (2,20.9) | 10.6 (3.5,20.6) | 3.2 (2.5,18.6) | 0.29 | 0.29 |
| **Clinical Outcomes** |  |  |  |  |  |  |
| Diabetes Mellitus | 12:8:1 Cases<br>108:59:4 Controls | 10% | 12% | 20% | 0.93 | 0.98 |
| Hypertension | 30:16:2 Cases<br>90:51:3 Controls | 25% | 24% | 40% | 0.91 | 0.61 |
| Myocardial Infarction | 2:6:2 Cases<br>118:112:3 Controls | 1.6% | 5.1% | 40% | 0.04 | 0.19 |

ALT, alanine aminotransferase; ALP, alkaline phosphatase; AST, aspartate aminotransferase; BIT, total bilirubin; BP, blood pressure; bpm, beats per minute; BMI, body mass index; cm, centimeter; dl, deciliter; eGFR, estimated glomerular filtration rate; fl, femtoliters; g, grams; HDL-C, high density lipoprotein cholesterol; HbA1c, hemoglobin A1C; kg, kilograms; L, liter; LDL-C, low density lipoprotein cholesterol; m, meters; MCH, mean corpuscular hemoglobin; MCHC, mean corpuscular hemoglobin concentration; MCV, mean corpuscular volume; mmHg, millimeters mercury; mg, milligrams; min, minutes; ml, milliliter; mmol, millimoles; ng, nanograms; pg, picograms; U, units; WHR, waist-to-hip ratio. All analyses were performed using linear mixed models with age and gender as fixed effects, and family ID as a random effect.

C211 is located in the mature form of GDF15 and forms a disulfide bridge with a more distal cysteine, C274, as part of a stabilizing network of bonds tying together beta strands (PDB 5VT2). We therefore predicted that C211G would destabilize the protein. To test the processing and stability of C211G, cDNA expression constructs were transiently transfected into GDF15-deficient HEK293 cells. By western blot, GDF15 WT reference allele was observed as full-length proprotein and processed mature protein in both cell lysate and supernatant (Figure 1B). GDF15 C211G, however, was observed only as full-length pro-protein retained in the cell lysate, indicating that this variant is expressed but neither processed nor secreted and therefore a true LOF variant. In a broader *in vitro* screen testing the functional necessity of all disulfide-forming cysteines in GDF15 (Figure 1C), we mutated cysteines to structurally analogous serine (Figure 1D). C273 was fully tolerant to substitution *in vitro* whereas C203 and C210 were partially tolerant (Figure 1D), indicating the susceptibility of GDF15 stability to disulfide bond disruption. The biological activity of each variant was tested *in vivo* through hydrodynamic injection (HDI) into WT mice. Consistent with *in vitro* expression data, GDF15 variants at positions 203, 210, and 273 exhibited body weight and food intake reductions comparable to WT GDF15 whereas other variants had little to no activity (Figure 1E). In total, these data indicate that mutations that disrupt C211 (or its bonding partner C274) result in loss of function both *in vitro* and *in vivo*.

### Burden analysis for LOF and C211G variants in PGR and UKBB

To analyze the effect of loss of function of GDF15 on a broad range of cardiometabolic phenotypes, we conducted a gene burden analysis in 75,018 individuals from PGR for all GDF15 LOF variants and the C211G missense variant collectively. Among 91 quantitative outcomes (Supplementary Table S1) and 6 binary outcomes (Supplementary Table S2) that were tested, we did not observe any significant associations at a false discovery rate (FDR) < 5% (Figure 2, Extended Data Table 2, and Supplementary Tables S1 and S2) under an additive model of inheritance. Waist-to-hip ratio (WHR) was nominally lower (p < 0.05). KOs had nominally significant longer QT intervals (P-recessive = 0.047) and fibroblast growth factor 23 (FGF23) concentrations, associated with arrhythmia^21^, were also trending higher (P-recessive = 0.058). In additive and recessive models, no associations were seen among the tested clinical outcomes of diabetes mellitus, atrial fibrillation/irregular heartbeats, myocardial infarction, angina, stroke, and atherosclerotic cardiovascular disease.

**Figure 2.**
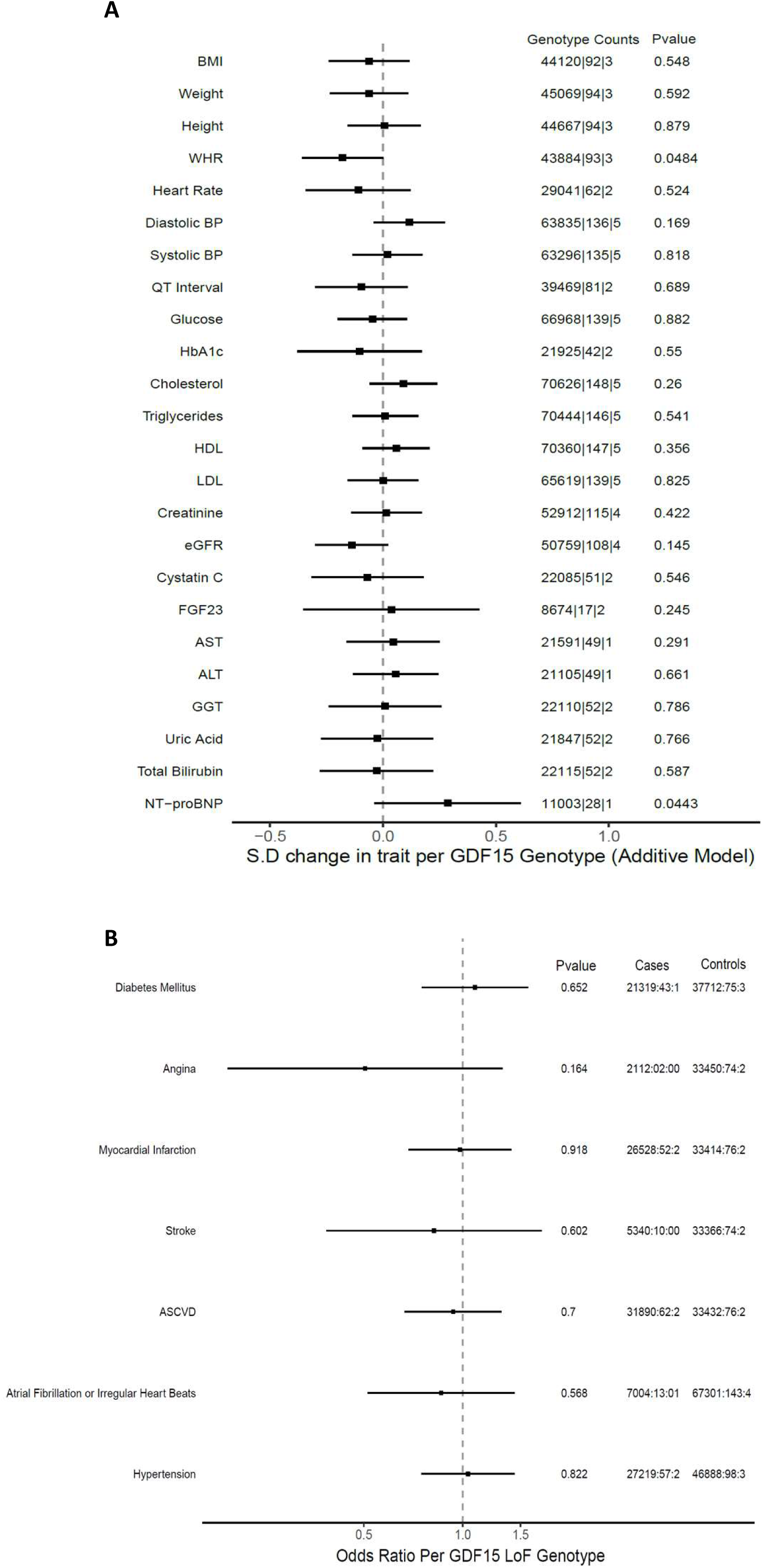
Forest plots, with 95% confidence intervals, of traits for GDF15 LOF and C211G carriers in PGR. (A) Quantitative traits. (B) Binary traits. Genotype counts for cases and controls are given as HomRR|HetRA|HomAA; R=reference allele; A=alternate allele. P-values are generated using whole genome regression models adjusting for age, sex, age*sex, age^2^ and top 10 genetic principal components (PCs). Additionally, Firth correction was used for binary traits. Quantitative effect estimates, per standard deviation (S.D.) units, are generated by standard linear regression adjusting for age, sex, age*sex, age^2^ and top 10 genetic PCs.

To see if any of the additive associations were consistent in another cohort, we identified 598 heterozygous carriers of LOF variants or C211G in the UK Biobank (UKBB) (Extended Data Table 3). We analyzed 25 phenotypes which overlapped with the PGR and meta analyzed the results. WHR did not replicate in the UKBB and we did not observe any other significant association at an FDR < 5% (Extended Data Table 4).

### Clinical characterization of GDF15 LOF and C211G carriers through recall-by-genotype (RBG) studies

For deeper clinical phenotyping of GDF15 LOF carriers, a recall-by-genotype (RBG) study was initiated for carriers of either GDF15 LOF or C211G variants. Carriers were re-contacted and consenting individuals and family members were enrolled in the study. All participants were Sanger-sequenced to ascertain variant carrier status (see Figure 3A for a sample partial pedigree of C211G carriers with corresponding Sanger sequencing results). In total, 192 participants were enrolled from 12 different families: 120 non-carriers, 67 heterozygous carriers, and 6 homozygous carriers of which 3 were identified through RBG studies (Extended Data Table 5).

**Figure 3.**
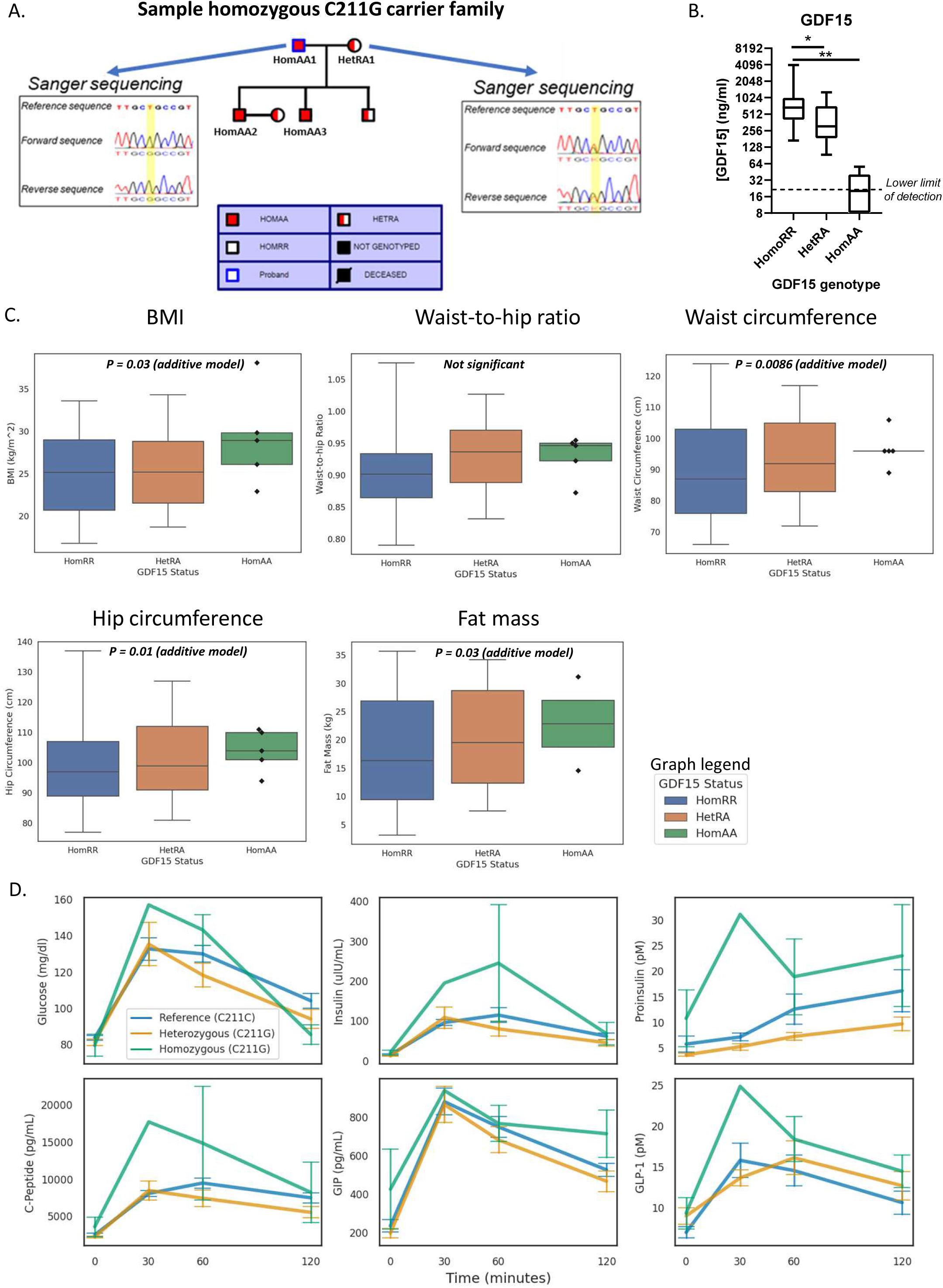
Profiling of GDF15 LOF and C211G carriers in PGR and recall-by-genotype (RBG) studies. (A) Partial pedigree of C211G carrier family from RBG study. GDF15 carrier couple, including a knockout, has >3 children. Their son, HomAA3, has multiple children (data not shown). (B) Circulating GDF15 in LOF heterozygotes and C211G heterozygotes and homozygotes. Homozygous carriers are near or below the lower limit of assay detection. Statistical analyses conducted by one-way ANOVA with Tukey’s multiple comparisons test. *p<0.05, **p<0.01. (C) BMI, waist-to-hip ratio, waist circumference, hip circumference, and fat mass by GDF15 status in the RBG study. Individual points are shown for homozygous LOF carriers only. Lower and upper whiskers represent the 5^th^ and 95^th^ percentiles respectively. All traits were modeled using linear mixed models with age and gender as fixed effects and family ID as a random effect. Sample sizes are shown as HomRR|HetRA|HomAA; R=reference allele; A=alternate allele. (D) Oral glucose tolerance test (OGTT) data from RBG studies. Data shown are from non-diabetic individuals identified in the RBG study. Sample sizes for the 0-minute (fasted), 60-minute, and 120-minute timepoints are shown as HomRR|HetRA|HomAA; R=reference allele; A=alternate allele. The 30-minute timepoint was assessed in only a subset of individuals: 25 non-carriers and 7 C211G carriers.

To confirm loss of circulating GDF15 protein in LOF variant carriers, GDF15 concentration was measured by ELISA (Figure 3B). LOF and C211G variants showed gene-dosage dependent reduction in circulating GDF15 concentration. Heterozygous carriers had roughly half the concentration of homozygous reference carriers and homozygous C211G carriers had concentrations that were below the limit of detection, though we cannot exclude the possibility that the ELISA detects an epitope lost by a change in C211.

Baseline characteristics of carriers including anthropometric traits, blood clinical chemistry tests, lipid panels, complete blood cell differential percentages, and disease history for RBG participants are shown by genotype in Table 1. A phenome-wide association study (PheWAS) of this data did not detect any significant associations at an FDR of 5% (Table 1). In an additive regression model, several nominal and weak associations were observed (Table 1, Figure 3C): higher BMI (P=0.03), higher weight (P=0.03), higher fat mass (P=0.03), higher waist circumference (P=8.6E-3), higher hip circumference (P=0.01), higher total cholesterol (P=0.04), and higher rate of myocardial infarction (P=0.04). The observation of higher incidence of MI may be driven by ascertainment bias since a subset of GDF15 LOF carrier probands were recruited from an MI case cohort within PGR. In a recessive model, several nominal associations were observed: lower HDL-C (P=0.03), lower LDL-C (P=0.04), and higher triglycerides (P=0.05). However, these nominal associations were driven by KOs from a single family in which several closely related non-KOs were also observed to have very high triglycerides and low LDL-C and HDL-C, suggesting a cause independent of GDF15 genotype. Oral glucose tolerance tests (OGTT) were also performed on a subset of RBG participants (Figure 3D and Extended Data Figure). Glucose, insulin, proinsulin, C-peptide, gastric inhibitory polypeptide (GIP), and glucagon-like peptide 1 (GLP1) were measured. No significant genotype-dependent changes in these parameters were observed.

Several GDF15 KOs exceeded the median life expectancy (66 years) in Pakistan, showed no overt enrichment of specific diseases, and have had children. A carrier couple, comprising male KO HomAA1 married to a female heterozygous C211G carrier HetRA1 (partial pedigree shown in Figure 3A), has successfully had multiple children, two of whom were GDF15 KO males (HomAA2 and HomAA3). One of these KO offspring (HomAA3) has also had multiple children (data not shown). HetRA1 did not report any nausea or morning sickness across any of her pregnancies. We also identified a GDF15 KO mother who has had multiple children, including a confirmed C211G heterozygous carrier.

## Discussion

We employed human genetics to explore the physiological impact and therapeutic implications of partial and complete GDF15 functional loss. In PGR, we identified GDF15 LOF variant carriers, including a homozygous carrier (KO). We also identified heterozygous and homozygous carriers of GDF15 missense variant C211G, a missense variant published previously^3^, and confirmed here, to be a functional LOF. We identified additional heterozygous and homozygous carriers of LOF and C211G variants in a RBG study accompanied by recruitment of family members. Cumulatively in PGR whole exomes and the follow-up RBG study, we identified a total of 8 homozygous LOF carriers ranging in age from 31 to 75 years. The carriers showed no consistent, overt metabolic dysfunction. This study represents the largest cohort of confirmed heterozygous GDF15 LOF carriers; 67 total vs the 11 previously described^3^.

GDF15 functional loss was nominally associated with higher BMI, weight, fat mass, waist circumference, hip circumference, total cholesterol, and rate of myocardial infarction. These associations were not significant in meta-analysis of PGR with UKBB. The observed trends for BMI and fat mass are consistent with preclinical studies but the magnitude of change is minimal. In GDF15 LOF and C211G variant carriers relative to non-carriers, OGTT data indicated no differences in concentration of glucose, insulin, proinsulin, C-peptide, GIP, or GLP1. There was no association between GDF15 carrier status and the risk of type 2 diabetes or hemoglobin A1c (HbA1c) concentration. In aggregate, these observations suggest that GDF15 LOF does not impact glucose metabolism.

GDF15 agonism has been tested in the clinic^2^, motivated by observations in pre-clinical models linking GDF15 to body weight and cachexia. The minimal impact of GDF15 LOF carrier status on BMI or metabolic phenotypes is consistent with clinical trial data showing that GDF15 agonist administration reduced body weight only modestly in obese participants^2^. Studies of naturally occurring human GDF15 gain-of-function variants, were they to be identified, would more rigorously test the hypothesis that GDF15 agonists reduce body weight.

The association of GDF15 with cachexia and mortality across multiple disorders has prompted the development of GDF15 antagonists for several indications. However, the pathophysiological consequences of inhibiting GDF15 in humans long-term is unclear. Catalym is testing visugromab in Phase 1 (clinicaltrials.gov ID NCT04725474) and Phase 2 (NCT06059547) trials to treat solid tumors. Aveo Oncology is testing AV-380 in Phase 1 trials to treat patients with cancer cachexia (NCT05865535). Pfizer is testing ponsegromab in Phase 2 trials in patients with cancer cachexia (NCT05546476) or heart failure (NCT05492500). The existence of GDF15 KOs suggests that pharmacological inhibition of GDF15 may be tolerated in non-pregnant adults.

Potential inhibition of GDF15 for the treatment of hyperemesis gravidarum^3^ is confounded by the natural increase of GDF15 during pregnancy. This elevation suggests a biological function during fetal development. Exploring this biology in rodent models is challenging as it appears to be a largely primate-specific phenomenon^22^. GDF15 LOF carriers, including KOs, report having had children, indicating that complete functional genetic loss of GDF15 in at least one parent is compatible with life and fertility. The existence of both male and female GDF15-null individuals who reached adulthood and had children suggests that reduction of GDF15 did not cause long-term impact on fertility, the mother, or the fetus. These observations raise the possibility that some degree of GDF15 inhibition, for example to treat anorexia or cachexia, may be tolerated, with the caveat that pharmacological intervention is not equivalent to genetic variation. The existence of human GDF15 KOs provides an opportunity to understand further the biological function of this factor during pregnancy.

## Table legends

*Supplementary Table S1*. Summary statistics for all 91 quantitative traits association with GDF15 LOFs and C211G gene burden under an additive model of inheritance.

*Supplementary Table S2*. Summary statistics for 6 clinical outcomes and binary traits association with GDF15 LOFs and C211G gene burden under an additive model of inheritance.

## Methods

### Whole exome sequencing studies

*Ethics statement.* The Institutional Review Board (IRB) at the Center for Non-Communicable Diseases (IRB: 00007048, IORG0005843, FWAS00014490) approved the study. This study has also been approved by the National Bioethics Committee for Research Pakistan (Reference number: NBC-756). All participants gave written informed consent.

*PGR variant QC and annotation.* Exome sequencing was performed at two different locations, at the Broad institute as described earlier^8^ and at the Regeneron Genetics Center^23^. All samples were sequenced at 30X coverage. Samples with low allele balance (<0.2) or low depth (<10) were set to missing and variants which had a missing rate >5% were removed. High confidence LOF variants were annotated as stop gained, frameshift, splice donor, and splice acceptor variants affecting more than 5% of the protein.

*PGR exome analysis.* All quantitative traits were transformed by the rank based inverse standard normal function, applied within each genotyping batch. Quantitative traits were analyzed using linear regression as implemented in regenie^24^. All analyses were adjusted for age, sex, age*sex, age^2^ and the top 10 genetics principal components (PCs) generated using common genotyping array SNPs. For exomes data, if genotyping array data wasn’t available, PCs were derived from common (MAF > 1%) exome SNPs. Exomes and genomes data were analyzed separately across sequencing centers and meta-analyzed using inverse variance weighted meta-analysis as implemented in METAL^25^. Binary traits were analyzed using a logistic regression model, with Firth fallback. Phenotypes were used only if 5 or more carriers were available for the trait in question.

*Phenotype definitions.* LDL was calculated using the Friedwald equation. eGFR was calculated using the CKD-EPI calculation. The LDL analysis was subset to individuals who were not on cholesterol lowering drugs, glucose concentrations were analyzed for participants who were not on oral antidiabetic drugs, and creatinine and eGFR were subset to participants without heart failure. Myocardial infarction cases were enrolled at time of event as described^9^. Type 2 diabetes was defined as individuals having an HbA1c >=6.5 or self-reporting to have diabetes or using oral-hypoglycemics. Individuals with a diabetes age of onset <30 years were excluded from the analysis. Type 2 diabetes controls were ascertained as individuals with HbA1c <6.5 or no self-reported history of diabetes and random blood glucose concentrations <150 mg/dl. Stroke cases were enrolled either at time of the event or were self-reported. Angina, atrial fibrillation/irregular heartbeats, and hypertension were all self-reported. Atherosclerotic cardiovascular disease cases were defined as any individuals with myocardial infarction, stroke, or angina. Controls were healthy individuals without cardiovascular disease history.

*UK Biobank Exome analysis.* UK Biobank analysis was performed in a similar manner to the PGR PheWAS analysis. Analyses were restricted to a set of unrelated individuals with high quality genetic data (UKBB Data Field 22020). A quality-controlled set of genotyping array data (UKBB Data Field 22418) was used in regenie’s whole genome regression step 1. All quantitative traits were transformed by the rank based inverse standard normal function. Quantitative traits were analyzed using linear regression as implemented in regenie^24^. All analyses were adjusted for age, sex, age*sex, age ^ 2 and the top 10 genetics PCs generated using common genotyping array SNPs. Binary traits were analyzed using a logistic regression model with Firth fallback.

*Exome meta-analysis.* Common phenotypes available in UKBB and PGR for GDF15 C211G missense and LOF variants were meta-analyzed using METAL^25^.

### *In vitro* GDF15 variant expression and western blotting

*Construct design.* Human codon-optimized GDF15 cDNAs were generated by gene synthesis in mammalian expression vector pcDNA3.1(+) to encode a CD33 signal peptide (PLLLLLPLLWAGALA) followed by an N-terminal 6x polyhistidine tag, residues 30-308 of human wild-type (NCBI accession no. NP_004855) or mutant proGDF15, and a C-terminal amyloid precursor protein (APP) epitope tag (EFRHDSG).

*Protein expression and extraction.* Proteins were expressed in suspension-adapted HEK293F or Expi293 cells. Cells were grown in a 37 degree C shaking incubator to a density of 1×10^6^/mL in serum-free media (FreeStyle or Expi293 media, Thermo Fisher) and transiently transfected using polyethylenimine (PEI 25K, Polysciences). After 72 hours, transfected cells were harvested by centrifugation at 500xg for 5 min. Conditioned media was collected for analysis by Western Blot. Cell pellets were washed with PBS, then resuspended in RIPA lysis and extraction buffer supplemented with 1X Halt protease inhibitor cocktail (ThermoFisher). Clarified cell lysate was collected by centrifugation at 16,000xg for 15 min and analyzed by Western Blot.

*Western blot.* Proteins in media and lysate samples were separated by SDS-PAGE and transferred to polyvinylidene fluoride (PVDF) membranes. Blots were probed with a primary antibody against the C-terminal APP tag followed by horseradish peroxidase (HRP)-conjugated secondary antibody to detect both proGDF15 and mature GDF15.

### In vivo studies

*Statement on animal welfare.* All animal work was performed under protocols reviewed and approved by the Novartis Institutes for BioMedical Research (NIBR) Animal Care and Use Committee. All procedures in these studies were in compliance with the Animal Welfare Act Regulations 9 CFR Parts 1, 2 and 3, and other guidelines (Guide for the Care and Use of Laboratory Animals, 2011).

*Animals, maintenance conditions, and diet.* Male diet induced obese (DIO) mice (C57BL/6NTac) fed a 60% fat diet (Research Diets D12492i) from 6-weeks of age onward were purchased from Taconic. Upon arrival, mice were housed one animal per cage under a 12-hour:12-hour light-dark cycle and received a minimum of 1 week acclimation prior to any use.

*Hydrodynamic DNA injections.* On the day of study, mice were placed in fresh cages, and the old food removed. Each study animal received a single hydrodynamic injection of plasmid DNA via tail vein. DNA (3 micrograms/mouse; pLEV113; LakePharma, Belmont, CA) encoding wild-type full-length human GDF15 or full-length human GDF15 with the conserved cysteine residues mutated to serine as indicated was diluted in sterile saline at a volume ∼7% of the animal’s body weight and rapidly injected within ∼5-10 seconds. Immediately after injection, pre-weighed fresh high-fat diet was added to each cage and animals returned to housing racks. Food intake and body weight were measured at the indicated time points.

### Recall-by-genotype study

*Overview.* Carriers of GDF15 variants were contacted by the Center of Non-Communicable Diseases in Karachi Pakistan under the IRB approval of IRB committee of Center for Non-Communicable Diseases (NIH registered IRB 00007048). After obtaining consent from the proband and from the family members, questionnaires regarding past medical and family history were administered by trained research staff, in the local language. Physical measurements like height, weight, waist and hip circumference were taken in standing position by using height and weight scales and a spring gauge tape measure. Blood pressure and heart rate were recorded by using OMRON healthcare M2 blood pressure monitors. Non-fasting blood samples were collected from each participant in EDTA and Gel Tubes. Serum and plasma were separated within 45 minutes of venipuncture. A random urine sample was also collected from each participant. The samples were stored temporarily in dry ice in the field and transported to a central laboratory based at CNCD and stored at –80 degrees Celsius. Measurements for total-cholesterol, HDL cholesterol, LDL cholesterol, triglycerides, VLDL, AST, ALT and creatinine were made in serum samples using enzymatic assays, whereas concentrations of HbA1c were measured using a turbidimetric assay in whole-blood samples (Roche Diagnostics, USA). Urinary microalbumin was also measured through the urine sample.

*Isolation of genomic DNA*. DNA was extracted from leukocytes of peripheral whole blood using a reference salting out method. DNA concentrations were determined by UV based quantification by using Nanodrop™ 2000 Spectrophotometer (Thermo Scientific™, USA).

*Sanger sequencing.* Whole blood-derived DNA from recalled participants was used to assess carrier status and zygosity for variants of interest via Sanger sequencing. Sanger sequencing was conducted at Macrogen, Inc (South Korea) or in-house at the CNCD. For Macrogen-processed samples, DNA samples were shipped directly to Macrogen for PCR amplification and Sanger sequencing. PCR primers were designed covering a region of approximately 200 to 300 bases around the variant. For in-house Sanger sequencing, specific primers were designed to amplify the region of interest using Platinum Master Mix (Thermo Scientific™, USA). This amplified DNA product was cleaned up using ExoSAP-IT Express PCR Product Cleanup (Thermo Scientific™, USA). The DNA was then used in BigDye Terminator v3.1 cycle sequencing following addition of BigDye XTerminator (Thermo Scientific™, USA) for cleanup and run on Applied Biosystems SeqStudio Genetic Analyzer (Thermo Scientific™, USA). Manufacturers’ protocols were followed for all kits.

*Oral glucose tolerance test (OGTT) protocol*. Consenting participants were requested to be on a high carbohydrate 3 days prior to testing. Blood samples from 8 hour fasting participants were obtained. Participants were then given a glucose drink (75g, 7.5 oz) and 3ml blood was collected on the 0, 30, 60 and 120-minute time interval. For each sample plasma and serum was separated, the samples were stored on dry ice and transferred to a –80C central location whereafter glucose concentrations were measured for each time point.

*GDF15 ELISA measurements.* GDF15 concentrations in human serum and plasma samples were quantified by ELISA using a commercially available kit (R&D cat# DY957) with some modifications to the kit method. Capture antibody was coated in PBS, 2μg/ml, 20μl/well overnight in a 384-well clear Maxisorp plate (Thermo cat# 464718). The plate was washed three times using wash buffer (PBS, 0.05% Tween-20), 100μl/well with a platewasher (Biotek). The plate was blocked with blocking buffer (PBS, 5% BSA (Fraction V fatty-acid free, EMD Millipore cat# 126575), 0.05% Tween-20, 0.05% TritonX-100), 75μl/well, 1hr. A standard curve was prepared in assay buffer (PBS, 2% BSA, 0.05% Tween-20, 0.05% TritonX-100) starting at 2ng/ml and serially diluting 1:3 for seven points plus a 0ng/ml control. Unknown samples were titrated in assay buffer starting at 1:2 (50% sample) and serially diluting 1:2 for four points. The ELISA plate was washed and samples were added in duplicate, 20μl/well, 1hr. The plate was washed and secondary antibody was added at 12.5ng/ml in assay buffer, 20μl/well, 1hr. The plate was washed and detection Streptavidin-HRP (Thermo cat# N200) was added at 1:10,000 in assay buffer, 20μl/well, 30min. The plate was washed and developed with TMB (Thermo cat# 34029), 20μl/well, 7min. The developing reaction was stopped with 2N sulfuric acid, 10μl/well and the plate was read on a Spectramax M5e at absorbance 450-570. Data were analyzed with SoftmaxPro v5.4.1. GDF15 concentration in unknown samples was determined by interpolation from the standard curve only for dilutions that fell on the linear range of the standard curve.

*MSD assays.* Circulating concentration of insulin, GLP-1 (total), GIP (total), C-peptide and proinsulin were evaluated using multiplexed Meso Scale Discovery (MSD) U-Plex Metabolic Group 1 assay plates according to manufacturer instructions (MSD, catalog no. K151ACL-2). Briefly, 96-well multiplex plates were coated 50 μL per well with biotinylated antibodies coupled to site-specific U-Plex linkers, allowing each analyte to self-assemble on unique spots in each well. Plates were incubated overnight at 4 degrees C with shaking at 500 rpm. Each well was washed three times with 150 µL of wash buffer (provided in kit) before the addition of 50 µL of recombinant protein or human plasma diluted in metabolic assay working solution (provided in kit). Plasma was diluted 8-fold for measurements of GIP, GLP-1, C-peptide, and insulin measurements, or 4-fold for proinsulin measurements. After shaking at 500 rpm for 2 h at room temperature, plates were washed three times with 150 µL wash buffer, treated with 50 µL of detection antibodies conjugated to electrochemiluminescent SULFO-TAG labels per well, and incubated at room temperature for 1hr with shaking. Following three washes with wash buffer,150 µL of MSD-GOLD read buffer B was added to each well, and plates were imaged on a MESO Sector S 600MM Imager (Meso Scale Discovery).The lower limits of quantification (LLOQ) for each analyte were in line with kit specifications (insulin: 0.18 µU/mL; pro-insulin: 1.97 pM; C-peptide: 110 pg/mL; GIP total: 3 pM; and, GLP-1 total: 54.3 pg/mL)

*Recall-by-genotype PheWAS.* PheWAS for quantitative traits in the recall study was performed using a linear mixed model with age and sex as fixed effects, and family ID as a random effect. All quantitative traits were transformed by the rank based inverse standard normal function. C211G carriers and carriers of LOF variants were analyzed jointly through burden tests. Both additive and recessive models were considered for all traits. Binary traits were analyzed using logistic regression with age and gender as covariates. All analyses were performed using the statsmodel Python module (version 0.13.0)^26^.

*Recall-by-genotype oral glucose tolerance test association analysis.* For each measurement (glucose, insulin, proinsulin, c-peptide, GIP, and GLP1), each time point as well as area under the curve were analyzed using a linear mixed model with age and sex as fixed effects, and family ID as a random effect. Distributions were transformed by the rank based inverse standard normal function prior to analysis. Both additive and recessive models were used for all traits. Analyses were restricted to individuals meeting the following criteria: no reported history of diabetes, HbA1c <6.5%, and no reported use of glucose lowering medication.

## Data Availability

All academic requests to access relevant data should be sent to. CNCD will ask relevant investigators to sign a data confidentiality agreement which would limit any investigator not to de-identify any of the study participants.

## Acknowledgements

This research has been conducted using the UK Biobank Resource under Application Number 59456. D.S. has received funding from Regeneron Pharmaceuticals, Eli Lilly & Company, Novartis, Merck, Astra Zeneca, NGM Biopharmaceuticals Inc., GSK, Astellas Pharma Inc., and Novo Nordisk. Sequencing for 37,804 exomes was performed at the Regeneron Genetics Center, Tarrytown, New York. A.M.G., C.K., L.B.L., E.D., A.M.C., R.Z., M.E.C., Z.C, L.D.L, Y.H.C., R.S.S., D.P.D., A.B.G., and J.E.D. are employees of Novartis. B.D. is an employee of Tango Therapeutics. A.A. and J.L.H. are employees of AstraZeneca. K.T., M.A., and I.K. are employees of Astellas Pharma. A.R.S. and J.L.R. are employees of the Regeneron Genetics Center. We thank all study participants of the PGR and UK Biobank for their vital contributions.

---

**Extended Data Table 1.** List of GDF15 LOF variants identified in 68,203 exomes and 6,815 genomes in PGR.

| Chr:Pos:Ref:Alt<br>(GRCh38) | Rsid | Allele<br>frequency | Heterozygous<br>carriers | Homozygous<br>carriers | Variant<br>effect | HGVSp |
| --- | --- | --- | --- | --- | --- | --- |
| 19:18386195:C:CG |  | 6.6E-06 | 1 | 0 | frameshift | Q4AfsTer295 |
| 19:18386315:AC:A |  | 6.6E-06 | 1 | 0 | frameshift | S44QfsTer21 |
| 19:18386320:C:CA |  | 6.6E-06 | 1 | 0 | frameshift | E45RfsTer254 |
| 19:18386320:CAG:C |  | 6.6E-06 | 1 | 0 | frameshift | E45VfsTer253 |
| 19:18386394:GC:G |  | 6.6E-06 | 1 | 0 | frameshift | N70TfsTer68 |
| 19:18388382:TC:T |  | 6.6E-06 | 1 | 0 | frameshift | R126GfsTer12 |
| 19:18388412:G:A |  | 6.6E-06 | 1 | 0 | stop<br>gained | W135Ter |
| 19:18388423:C:T |  | 6.6E-06 | 1 | 0 | stop<br>gained | R139Ter |
| 19:18388517:T:TG |  | 6.6E-06 | 1 | 0 | frameshift | A171GfsTer128 |
| 19:18388559:T:TG |  | 6.6E-06 | 1 | 0 | frameshift | R185AfsTer114 |
| 19:18388567:C:T | rs552234112 | 0.00011 | 16 | 0 | stop<br>gained | Q187Ter |
| 19:18388575:CA:C | rs767520639 | 6.6E-06 | 1 | 0 | frameshift | R190GfsTer48 |
| 19:18388691:G:A | rs775018710 | 8.6E-05 | 11 | 1 | stop<br>gained | W228Ter |
| **All LOFs** |  | **2.7E-04** | **38** | **1** | **All LOFs** |  |
| 19:18388639:T:G | rs372120002 | 0.00086 | 123 | 4 | missense | C211G |
| **All LOFs + C211G** |  | **1.1E-03** | **161** | **5** | **All LOFs + C211G** |  |

Chr, chromosome; pos, position; Ref, reference allele; Alt, alternate allele; and HGVSp, Human Genome Variation Society protein level change.

**Extended Data Table 2.** PGR distribution of traits per GDF15 genotype status.

|  |  | HomRR | HetRA | HomAA |  |  |
| --- | --- | --- | --- | --- | --- | --- |
|  | Count | median (IQR)<br>or % | median (IQR) or % | median (IQR) or % | P-val<br>(additive) | P-val<br>(recessive) |
| Quantitative traits |  |  |  |  |  |  |
| Age | 74855:158:5 | 54 (47,60) | 55 (47,61) | 60 (60,65) |  |  |
| Gender (Female) | 20326:38:1 | 27% | 32% | 25% |  |  |
| BMI (kg/m<sup>2</sup>) * | 44120:92:3 | 26.5 (23.9, 29.3) | 26.1 (24.2, 28.5) | 26.8 (25.2, 27.1) | 0.548 | 0.86 |
| Weight (kg) * | 45069:94:3 | 70 (65.0, 80.0) | 70 (65.5, 78.8) | 70 (67.0, 77.5) | 0.592 | 0.88 |
| Height (m) * | 44667:94:3 | 165 (159.0, 170.0) | 165.5 (160.0, 169.8) | 165 (162.5, 171.5) | 0.879 | 0.97 |
| WHR * | 68792:143:5 | 0.97 (0.93, 1.01) | 0.96 (0.91, 1.0) | 0.97 (0.95, 1.01) | 0.023 | 0.31 |
| Heart Rate (bpm) | 29041:62:2 | 78 (70.0, 88.0) | 79.5 (71.3, 82.5) | 67 (62.5, 71.5) | 0.524 | 0.32 |
| BP diastolic (mmHg) | 63835:136:5 | 80 (76.0, 90.0) | 80 (80.0, 90.0) | 80 (80.0, 80.0) | 0.169 | 0.73 |
| BP systolic (mmHg) | 63296:135:5 | 130 (120, 140) | 129 (120, 140) | 130 (120, 130) | 0.818 | 0.65 |
| QT Interval (ms) | 39469:81:2 | 0.36 (0.32, 0.39) | 0.36 (0.32, 0.38) | 0.42 (0.41, 0.43) | 0.689 | 0.047 |
| Glucose (mg/dl) | 66968:139:5 | 118 (92.8, 173.0) | 115 (93.8, 166.4) | 117.4 (91.0, 139.4) | 0.882 | 0.69 |
| HbA1c (%) | 21925:42:2 | 6.3 (5.7, 7.7) | 6.2 (5.5, 7.0) | 6.1 (5.9, 6.2) | 0.551 | 0.48 |
| Cholesterol (mg/dl) | 70626:148:5 | 174 (143, 208) | 175 (148, 207) | 190 (135, 258) | 0.260 | 0.31 |
| Triglycerides (mg/dl) | 70444:146:5 | 157 (110, 229) | 168 (108, 225) | 146 (87, 356) | 0.541 | 0.77 |
| HDL-C (mg/dl) | 70360:147:5 | 34 (28, 41) | 34 (27, 42) | 45 (33, 48) | 0.356 | 0.14 |
| LDL-C (mg/dl) | 65619:139:5 | 104 (79, 132) | 104 (73, 128) | 77 (63, 156) | 0.825 | 0.84 |
| Creatinine (mg/dl) | 52912:115:4 | 0.89 (0.7, 1.1) | 0.9 (0.7, 1.1) | 0.95 (0.88, 1.1) | 0.422 | 0.41 |
| eGFR (ml/min/1.73m<sup>2</sup>) | 50759:108:4 | 87 (70, 105) | 84 (71, 101) | 70 (64, 81) | 0.145 | 0.10 |
| Cystatin C (mg/dl) | 22085:51:2 | 1 (0.8, 1.2) | 1 (0.8, 1.2) | 1.1 (1.0, 1.2) | 0.546 | 0.30 |
| FGF23 (pg/ml) | 8674:17:2 | 69 (50, 104) | 80 (53, 118) | 204 (184, 223) | 0.245 | 0.058 |
| GGT (U/L) | 22110:52:2 | 22 (15.0, 36.0) | 20.5 (13.8, 32.3) | 49.5 (40.3, 58.8) | 0.786 | 0.98 |
| Uric Acid (mg/dl) | 21847:52:2 | 5 (3.9, 6.0) | 4.75 (3.88, 5.7) | 5.7 (5.6, 5.8) | 0.766 | 0.41 |

| Total Bilirubin (mg/dl) | 22115:52:2 | 0.26 (0.16, 0.4) | 0.28 (0.18, 0.41) | 0.29 (0.23, 0.35) | 0.587 | 0.80 |
| --- | --- | --- | --- | --- | --- | --- |
| Binary Traits |  |  |  |  |  |  |
| Diabetes Mellitus | 21319:43:1 Cases<br>37712:75:3 Controls | 36% | 36% | 25% | 0.65 | 0.75 |
| Angina | 2112:2:0 Cases<br>33450:74:2 Controls | 5.9% | 2.6% | 0% | 0.16 | 0.86 |
| Myocardial Infarction | 26528:52:2 Cases<br>33414:76:2 Controls | 44.3% | 40.6% | 50% | 0.92 | 0.73 |
| Stroke | 5340:10:0 Cases<br>33366:74:2 Controls | 13.8% | 11.9% | 0% | 0.60 | 0.59 |
| Atherosclerotic Cardiovascular Disease | 31890:62:2 Cases<br>33432:76:2 Controls | 48.8% | 44.9% | 50% | 0.70 | 0.89 |
| Atrial Fibrillation or Irregular Heartbeats | 7004:13:1 Cases<br>67301:143:4 Controls | 9.4% | 8.3% | 20% | 0.57 | 0.34 |
| Hypertension | 27219:57:2 Cases<br>46888:98:3 Controls | 36.7% | 36.7% | 40% | 0.82 | 0.49 |

Hom, homozygote; Het, heterozygote; R, reference allele; A, alternate allele; IQR, interquartile range; ALT, alanine aminotransferase; AST, aspartate aminotransferase; BP, blood pressure; bpm, beats per minute; BMI, body mass index; dl, deciliters; eGFR, estimated glomerular filtration rate; GGT, gamma-glutamyl transferase; HbA1c, hemoglobin A1c; HDL-C, high density lipoprotein cholesterol; kg, kilograms; L, liters; LDL-C, low density lipoprotein cholesterol; m, meters; mg, milligrams; min, minutes; mmHg, millimeters mercury; ml, milliliters; ms, milliseconds; pg, picograms; U, units; WHR, waist-to-hip ratio. \*Individuals recruited at time of acute Myocardial infarction or stroke occurrence were removed. All analyses were adjusted for age, sex, age\*sex, age ^ 2 and the top 10 genetics PCs.

**Extended Data Table 3.** List of GDF15 LOF variants analyzed in UKBB.

| Chr:Pos:Ref:Alt (GRCh38) | Rsid | Allele frequency | Heterozygous Carriers | Variant effect | HGVSp |
| --- | --- | --- | --- | --- | --- |
| 19:18386267:T:TG | rs1971826619 | 7.70E-06 | 6 | frameshift | A29RfsTer270 |
| 19:18386279:GTC:G |  | 6.41E-06 | 5 | frameshift | L32GfsTer266 |
| 19:18386297:C:CCCGGGACCCT | rs777614840 | 5.13E-06 | 4 | frameshift | R37PfsTer265 |
| 19:18386349:C:T |  | 1.28E-06 | 1 | stop gained | R54Ter |
| 19:18386361:A:T |  | 5.13E-06 | 4 | stop gained | K58Ter |
| 19:18386369:C:G |  | 1.28E-06 | 1 | stop gained | Y60Ter |
| 19:18386409:G:T |  | 1.28E-06 | 1 | stop gained | E74Ter |
| 19:18386468:T:C | rs780075693 | 5.13E-06 | 4 | splice donor |  |
| 19:18388285:G:A |  | 1.28E-06 | 1 | splice acceptor |  |
| 19:18386452:TACTC:T | rs770376692 | 6.41E-06 | 5 | frameshift | L89RfsTer48 |
| 19:18388390:TC:T |  | 1.28E-06 | 1 | frameshift | P129RfsTer9 |
| 19:18388412:G:A |  | 1.28E-06 | 1 | stop gained | W135Ter |
| 19:18388413:G:A |  | 1.28E-06 | 1 | stop gained | W135Ter |
| 19:18388455:AC:A |  | 6.41E-06 | 5 | frameshift | Q151RfsTer87 |
| 19:18388459:C:T |  | 1.28E-06 | 1 | stop gained | Q151Ter |
| 19:18388567:C:T | rs552234112 | 1.15E-05 | 9 | stop gained | Q187Ter |
| 19:18388575:CA:C | rs767520639 | 6.93E-05 | 54 | frameshift | R190GfsTer48 |
| 19:18388576:AG:A | rs1254999181 | 1.28E-06 | 1 | frameshift | R192AfsTer46 |
| 19:18388614:CT:C |  | 1.28E-06 | 1 | frameshift | C203VfsTer35 |
| 19:18388638:CT:C |  | 1.28E-06 | 1 | frameshift | C211AfsTer27 |
| 19:18388683:G:A | rs2144616595 | 1.28E-06 | 1 | stop gained | W225Ter |
| 19:18388691:G:A | rs775018710 | 1.28E-06 | 1 | stop gained | W228Ter |
| 19:18388700:C:A | rs748621260 | 3.85E-06 | 3 | stop gained | S231Ter |
| 19:18388864:C:T |  | 1.28E-06 | 1 | stop gained | Q286Ter |
| **All LOFs** |  | **1.45E-04** | **113** | **All LOFs** |  |
| 19:18388639:T:G | rs372120002 | 622E-04 | 485 | missense | C211G |
| **All LOFs + C211G** |  | **7.67E-04** | **598** | **All LOFs + C211G** |  |

Chr, chromosome; pos, position; Ref, reference allele; Alt, alternate allele; and HGVS<sub>p</sub>, Human Genome Variation Society protein level change; LOF, loss-of-function.

**Extended Data Table 4.**
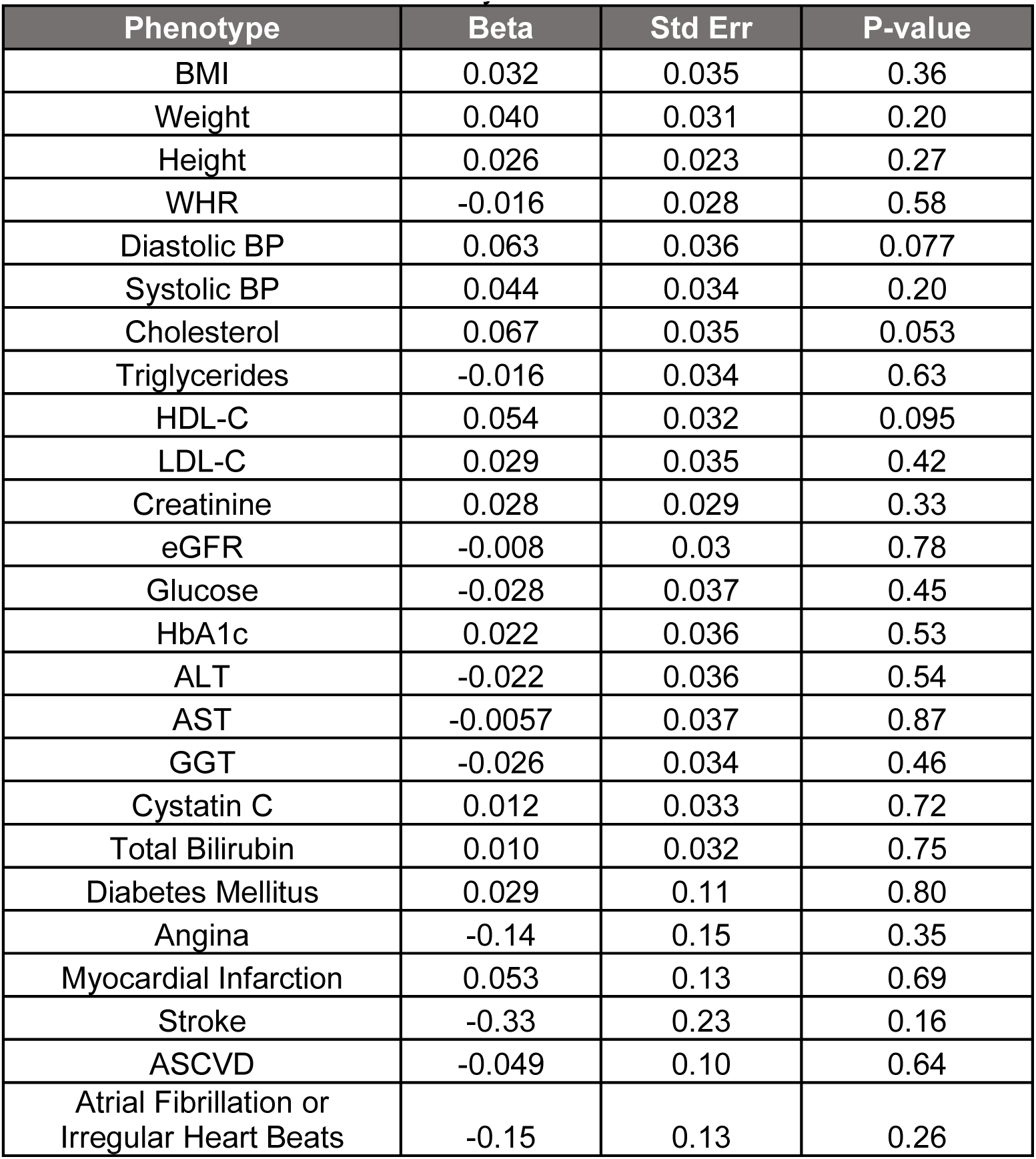
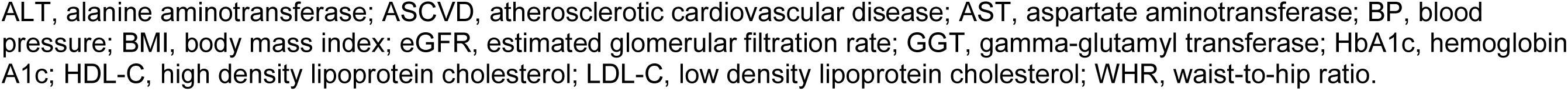
Meta-analyzed results of common traits from UKBB and PGR.

| Phenotype | Beta | Std Err | P-value |
| --- | --- | --- | --- |
| BMI | 0.032 | 0.035 | 0.36 |
| Weight | 0.040 | 0.031 | 0.20 |
| Height | 0.026 | 0.023 | 0.27 |
| WHR | -0.016 | 0.028 | 0.58 |
| Diastolic BP | 0.063 | 0.036 | 0.077 |
| Systolic BP | 0.044 | 0.034 | 0.20 |
| Cholesterol | 0.067 | 0.035 | 0.053 |
| Triglycerides | -0.016 | 0.034 | 0.63 |
| HDL-C | 0.054 | 0.032 | 0.095 |
| LDL-C | 0.029 | 0.035 | 0.42 |
| Creatinine | 0.028 | 0.029 | 0.33 |
| eGFR | -0.008 | 0.03 | 0.78 |
| Glucose | -0.028 | 0.037 | 0.45 |
| HbA1c | 0.022 | 0.036 | 0.53 |
| ALT | -0.022 | 0.036 | 0.54 |
| AST | -0.0057 | 0.037 | 0.87 |
| GGT | -0.026 | 0.034 | 0.46 |
| Cystatin C | 0.012 | 0.033 | 0.72 |
| Total Bilirubin | 0.010 | 0.032 | 0.75 |
| Diabetes Mellitus | 0.029 | 0.11 | 0.80 |
| Angina | -0.14 | 0.15 | 0.35 |
| Myocardial Infarction | 0.053 | 0.13 | 0.69 |
| Stroke | -0.33 | 0.23 | 0.16 |
| ASCVD | -0.049 | 0.10 | 0.64 |
| Atrial Fibrillation or<br>Irregular Heart Beats | -0.15 | 0.13 | 0.26 |

ALT, alanine aminotransferase; ASCVD, atherosclerotic cardiovascular disease; AST, aspartate aminotransferase; BP, blood pressure; BMI, body mass index; eGFR, estimated glomerular filtration rate; GGT, gamma-glutamyl transferase; HbA1c, hemoglobin A1c; HDL-C, high density lipoprotein cholesterol; LDL-C, low density lipoprotein cholesterol; WHR, waist-to-hip ratio.

**Extended Data Table 5.** GDF15 coding variants and corresponding carriers identified in the recall-by-genotype study and confirmed by Sanger sequencing.

| GRCh38 chr:pos | Reference allele | Alternate allele | Rsid | Variant effect | HGVSc | HGVSp | Non carriers | Heterozygous carriers | Homozygous carriers |
| --- | --- | --- | --- | --- | --- | --- | --- | --- | --- |
| 19:18386320 | C | CA |  | frameshift | c.132dup | E45RfsTer254 | 3<br>(0 probands) | 6<br>(1 proband) | 0 |
| 19:18388691 | G | A | rs775018710 | stop<br>gained | c.683G>A | W228Ter | 1<br>(0 probands) | 4<br>(1 proband) | 0 |
| 19:18388639 | T | G | rs372120002 | missense | c.631T>G | C211G | 116<br>(0 probands) | 57<br>(6 probands) | 6<br>(3 probands) |
| **All variants** |  |  |  |  |  |  | **120** | **67** | **6** |

Chr, chromosome; pos, position; HGVSc, Human Genome Variation Society coding level change; HGVSp, Human Genome Variation Society protein level change.

**Extended Data Figure.**
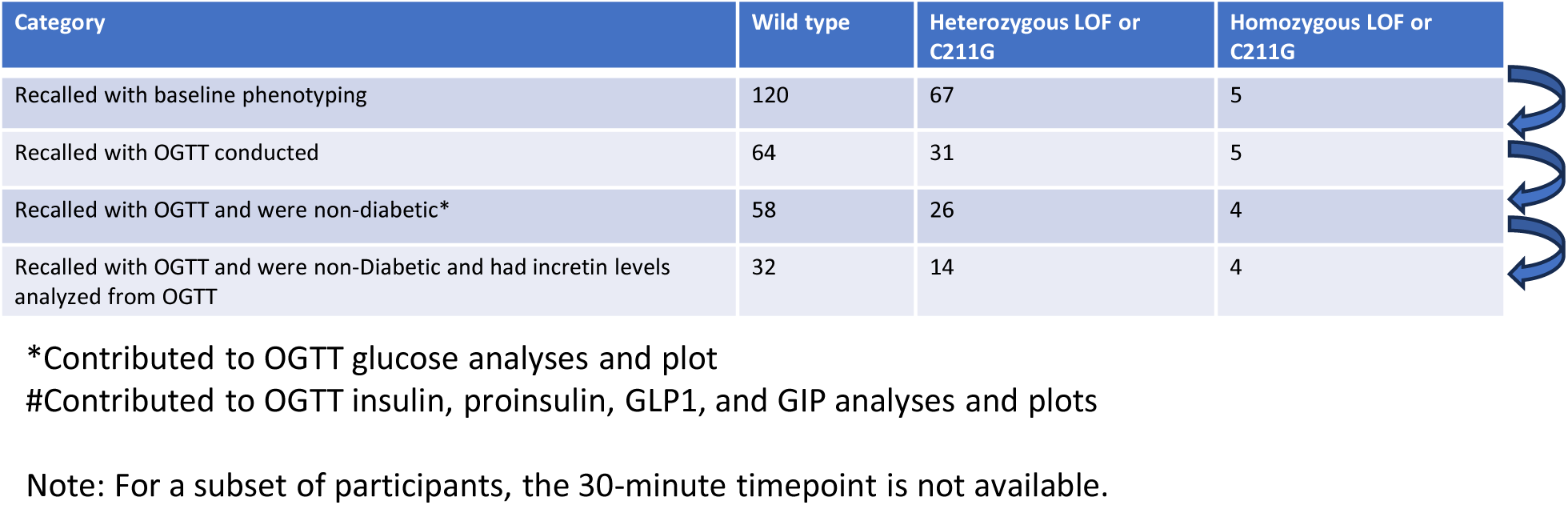
Breakdown of number of participants and corresponding applied phenotyping and analyses from the RBG study.

